# Association between adiposity and cardiovascular outcomes: an umbrella review and meta-analysis

**DOI:** 10.1101/2020.08.18.20176578

**Authors:** Min Seo Kim, Won Jun Kim, Amit V Khera, Hong-Hee Won

## Abstract

**Objective:** To evaluate the strength and certainty of the evidence underlying an association between increased adiposity, as assessed by body-mass index (BMI), waist circumference (WC), or waist-to-hip ratio (WHR) and identify the risk of incident cardiovascular disease (CVD) events or mortality

**Design:** Umbrella review of systematic reviews and meta-analyses.

**Data sources:** Google Scholar, PubMed, Embase, Cochrane Database of Systematic Reviews, and manual screening of retrieved references

**Eligibility criteria:** Systematic reviews or meta-analyses of observational studies and Mendelian randomisation (MR) studies that evaluated the association between various obesity-related indices and the risk of developing CVD and/or mortality due to CVD

**Data synthesis:** Eleven systematic reviews and 53 meta-analyses that investigated associations between obesity and cardiovascular outcomes were included. Results from recently published cohort studies were also incorporated into the existing meta-analyses to update them with more recent data. Thus, the present study compiled all the relevant evidence accumulated to date, encompassing a total of 488 cohorts and over 30 million participants. MR studies were collected to identify any causal relationship between obesity and various CVD outcomes, and to avoid reverse causality. The degree of obesity was measured with BMI, WC, and WHR. The evidence levels of pooled results were graded into high, moderate, low, and very low according to the Grading of Recommendations Assessment, Development and Evaluation framework.

**Results:** An increase in BMI was associated with a higher risk of developing coronary heart disease, heart failure, atrial fibrillation, stroke, hypertension, aortic valve stenosis, pulmonary embolism, and venous thromboembolism; the study results corroborate the casual effect of obesity on the incidence of CVD, except stroke, based on MR studies. The increase in the risk of developing CVD for every 5 kg/m^2^ increase in BMI ranged from 7% (relative risk [RR], 1.07; 95% confidence interval [CI], 1.03 to 1.11) for stroke to 49% (RR, 1.49; 95% CI, 1.41 to 1.58) for hypertension. The risk of all-cause mortality and CVD-specific mortality increased with adiposity, which was supported by a high grade of evidence from observational analyses; however, the causal effect of obesity on mortality outcomes was not significant in MR studies.

**Conclusions:** Only 15 out of the 53 associations (28%) reported for obesity and CVD outcomes were supported with high evidence levels from observational analyses. Although other reported associations might be valid, various degrees of uncertainty remain. The causal effect of obesity on 9 of the 14 CVD-related outcomes was corroborated by MR studies. As obesity is progressively increasing around the globe and because CVD remains a constant threat to public health, it is necessary to understand the gradient of evidence underlying the association between these two clinical entities. Any weak links in the association and causality discovered in this review should be reinforced with further scientific research, while high-certainty associations with established causality should be reflected in clinical practices and health policies.

**Systematic review registration:** PROSPERO CRD42020179469.

**Summary Box:** *Section 1:* What is already known on this topic - Obesity is progressively increasing worldwide and cardiovascular disease (CVD) remains a continued threat to public health.
- Although obesity as a risk factor for various cardiovascular outcomes has been studied for decades, the results from previous studies are heterogeneous, making it difficult for clinicians and policy makers to determine genuine and reliable associations.
- The level of evidence underlying the associations between obesity and CVD remains unknown.

*Section 2: What this study adds:*

- Only 15 of the 53 reported associations (28%) between obesity and CVD outcomes are supported with a high level of evidence. While other associations may be genuine, various degrees of uncertainty remain.
- An increase in body-mass index was associated with a higher risk of developing coronary heart disease, heart failure, atrial fibrillation, stroke, hypertension, aortic valve stenosis, pulmonary embolism, and venous thromboembolism; the casual effect of obesity on the incidence of CVD was corroborated by Mendelian randomisation (MR) studies, with the exception of stroke.
- The increase in the risk of all-cause mortality and CVD-specific mortality with adiposity was supported by a high grade of evidence in observational analyses, but the causal effect of obesity on mortality outcomes was not significant in MR studies.

## INTRODUCTION

Obesity has become a major public health challenge as cumulative evidence suggests that increased adiposity is a causative risk factor for diverse adverse health outcomes, including multiple cancers^1^, diabetes mellitus^2^, gynaecological-obstetric conditions^3^, and cardiovascular diseases (CVD)^4–6^. CVDs account for over two-thirds of deaths attributable to high body-mass index (BMI)^7^, and the consequential health outcomes constitute a major portion of health-related economic burden worldwide^8–12^. Despite countermeasures, the outlook is not favourable, and the incidence of CVD is expected to increase over the next few decades, especially in low- and middle-income countries, as the average BMI increases^13^.

Numerous studies support the association between obesity indices and cardiovascular outcomes, but considerable heterogeneity exists between these studies^14^. The heterogeneity may be attributable to the complex nature of obesity as a mediator-confounder. Obesity can directly impact diseases or syndromes, but is also subject to reverse causality, whereby the presence of disease may influence the BMI^15, 16^. Therefore, any investigation exploring this relationship must also address the direction of this causality. In addition, inherent biases are present in the study design of observational research, such as selection and publication biases, which can potentially inflate the observed effect^17–19^. A recent umbrella review revealed that despite strong claims of increased incidence of cancers with increased adiposity, only 11 (30%) of the 36 cancer types showed this association with reliable evidence, with minimal influence of biases^1^.

We conducted an umbrella review of systematic reviews and meta-analyses to appraise the entire context and quality of the vast amount of relevant evidence on the association between obesity indices and CVD. The results from recently published cohort studies were manually incorporated into existing meta-analyses to update previous results, and a total of 488 cohorts and over 30 million participants were integrated for quantitative syntheses. The collective outcomes were stratified into distinct evidence levels to quantify the level of certainty underlying each association and included Mendelian randomisation (MR) studies to minimise reverse causation bias. This work may help to contextualise the magnitude of the association and explicate the causality of obesity on CVD.

## METHODS

### Literature search and selection criteria

We systematically searched Google Scholar, PubMed, Embase, and the Cochrane Database of Systematic Reviews for systematic reviews and meta-analyses that investigated the association between adiposity indices and cardiovascular health outcomes, from inception to March 28, 2020. The adiposity indices of interest included BMI, waist circumference (WC), and waist-to-hip ratio (WHR). We used a predefined search strategy outlined in the supplementary appendix, and reference lists of relevant review articles were also screened to retrieve additional studies. Observational studies were also collected to update previous meta-analyses, and MR studies were incorporated to evaluate causality as has been performed in previous umbrella reviews^20, 21^. We imposed no restriction on language, but all included studies were written in English. The study protocol is published in PROSPERO (CRD42020179469).

### Inclusion and exclusion criteria

We included systematic reviews and meta-analyses of observational studies, as well as the MR studies that explored the association between obesity indices and cardiovascular outcomes using genetic instruments. We excluded systematic reviews and meta-analyses that evaluated indices other than BMI, WC, and WHR, such as weight loss %, history of bariatric surgery, and adipose tissue volume, as they can increase heterogeneity and hinder valid synthesis of results. Studies that included patients who underwent percutaneous coronary intervention or coronary artery bypass graft, while assessing the incidence of CVD, and those involving animal experiments or in-vitro results were also excluded.

When there was more than one meta-analysis study on the same topic, we preferentially included the most recent, or the one that comprised the largest number of studies to avoid duplication. For outdated meta-analyses, we incorporated the recently published cohort data into the meta-analysis, calculated the pooled effect sizes, and re-analysed the data for heterogeneity, publication bias, and prediction interval (Supplementary Table 1). When multiple observational or MR studies were conducted with an identical cohort (e.g., UK Biobank), those with the most available information (e.g., dose-response estimates) or those with the more comprehensively adjusted model was utilised for the update.

### Data extraction

Two researchers (MS Kim and WJ Kim) independently searched the existing literature and extracted the data. The titles, abstracts, and keywords of each study were reviewed for inclusion, and any ambiguity was resolved through discussion. The study selection process was recorded using the PRISMA flowchart^22^ (Figure 1).

**Fig. 1.**
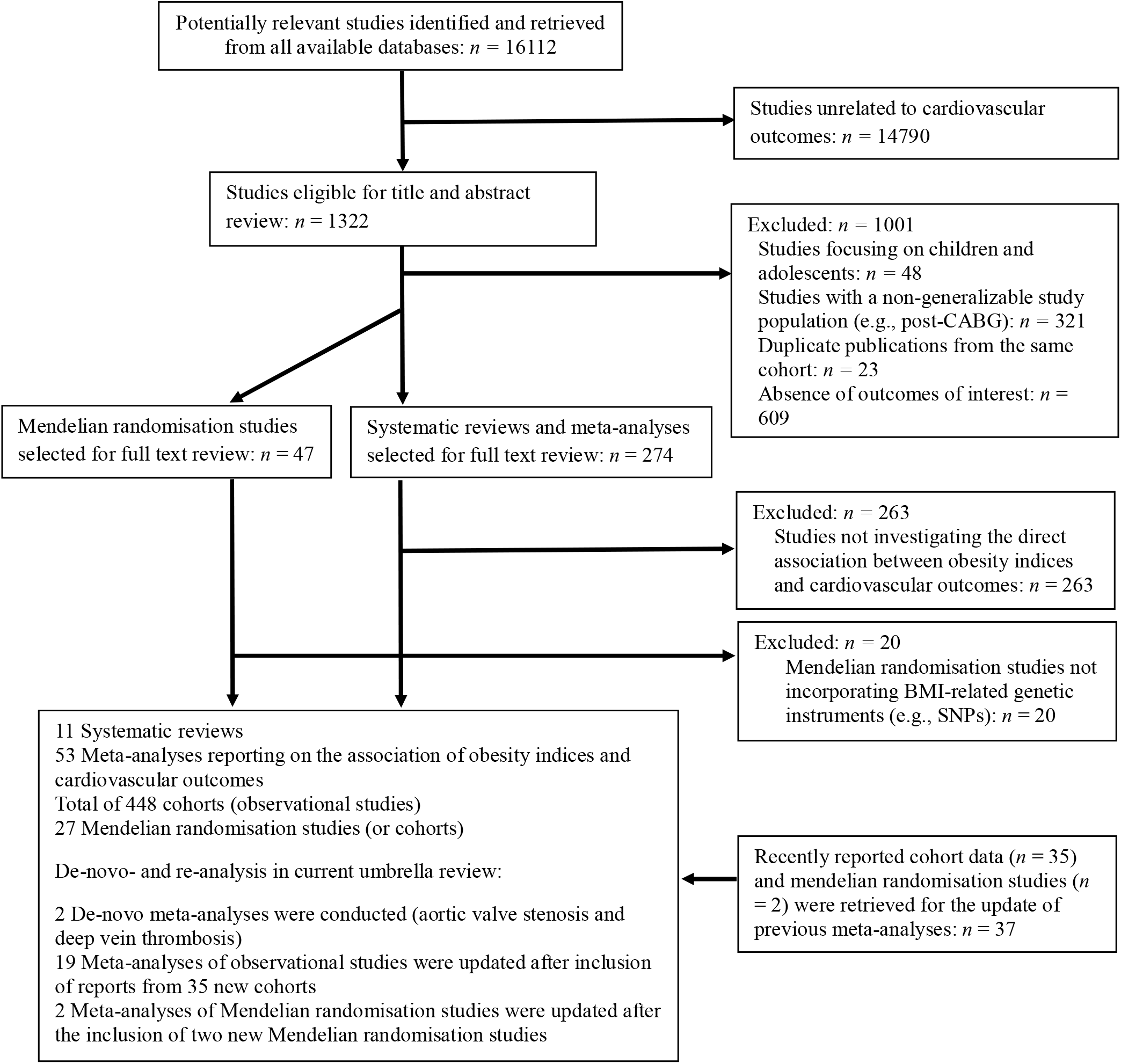
Flow diagram of the search and selection process

Data were collected with a predefined template. The following details were obtained from the included systematic reviews and meta-analyses of observational studies: publication year, number of studies included in the meta-analysis, exposures, comparisons, number of cases and participants, study design, model of effect estimation (random- or fixed-effects), heterogeneity, and maximally adjusted effect size with 95% confidence interval (CI) for each component study (Supplementary Tables 1 and 2). The adjustment factors included in the model were also retrieved to determine whether relevant confounders were accounted for. Both categorised (overweight, obese, and severely obese) and continuous (BMI, WC, and WHR) measures were extracted for qualitative synthesis. For MR studies, we extracted data on exposure, sample size, instrumental variable method, genetic instrument (GI), variance (R^2^) explained by GI, and maximally adjusted the effect estimates with 95% CI (Supplementary Table 3).

### Data analysis

We replicated the meta-analyses in our analytic framework and re-analysed the data to uncover the non-explicit details of these meta-analyses to evaluate the quality of evidence. The following items were considered: effect sizes in both fixed- and random-effects models; heterogeneity among studies that were calculated using I^2^ metric^23^; the presence of publication bias and small study effect using Egger’s tests (significance threshold, P < 0.10)^24^; p-curve test detecting p-hacking^25–28^; and 95% prediction intervals, representing the range within which the effect estimates of future studies will lie with 95% certainty^29–31^. We did not conduct the test for excess significance (TES) as it has not been thoroughly evaluated and is not currently recommended by the Cochrane Collaboration as an alternative to conventional tests of publication bias^32^; indeed, the TES was omitted in numerous previous umbrella reviews^33–37^.

We conducted a pairwise meta-analysis using R (version 3.6.0) software^38^ for re-analysis and update of previous meta-analyses with recently published observational studies. The results are reported in Supplementary Figures 2–39; further details of the methodology and our analytic workflow for pairwise meta-analysis are described elsewhere^39–42^. For certain phenotypes, such as aortic valve stenosis, that have not been meta-analysed despite a sufficient number of published original studies, we performed our own (de-novo) meta-analyses to pool the effect sizes and increase power. The summary of the effect estimation metric (odds ratio [OR], relative risk [RR], and hazard ratio [HR]) presented by each study is shown in Supplementary Table 1. To provide a straightforward comparison and synthesize multiple outcomes in a single visualization, as shown in Figures 2–4, we approximated different metrics to equivalent RRs using the guideline outlined by Fusar-Poli et al.,^43^ and adapted the approach of coordinating the results with different metrics used in previous meta-analyses^44 45^ and umbrella reviews^20 46^. Pre-specified subgroup analyses were performed to determine whether the results were affected by BMI categories or sex. We recalculated the dose-response relationship by pooling dose-response estimates from each study in the included meta-analyses if they were presented separately for BMI categories^47^.

**Fig. 2.**
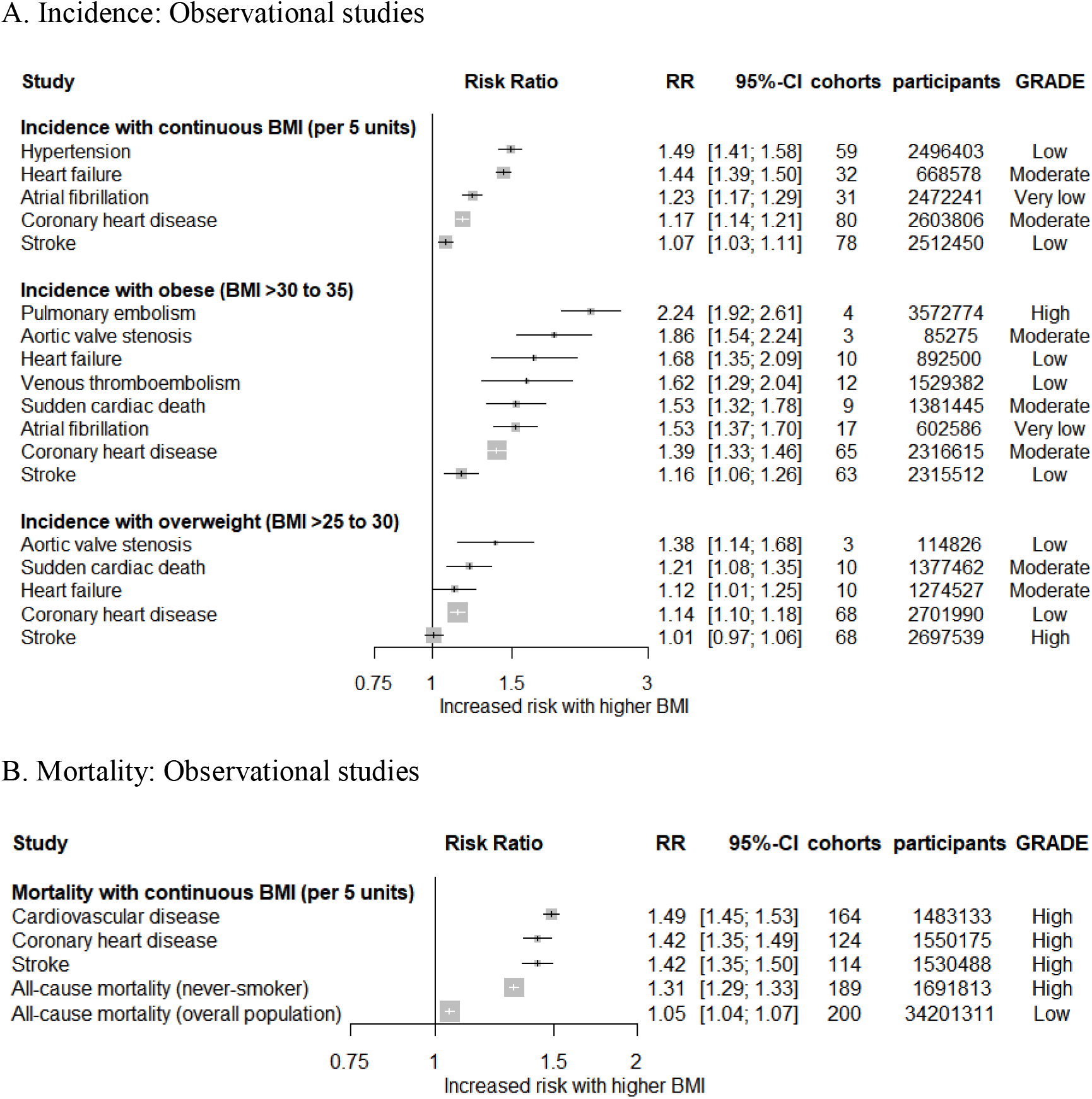
Collective results from observational studies. (A) Increased risk of cardiovascular events with elevated continuous and categorical BMI. (B) Increased risk of death with elevated continuous BMI. All results are based on random-effect models. The cohort and participant columns display the number of independent cohorts and the total number of participants incorporated in the meta-analysis for the outcome. The certainty of evidence underlying each association between BMI and cardiovascular outcomes was evaluated with the GRADE framework. GRADE, Grading of Recommendations Assessment, Development, and Evaluation; BMI, Body mass index; RR, Risk ratio or relative risk

The analytic process for MR was identical to that of the meta-analyses of observational studies. Since the reliability of the results of MR studies largely depends on the efficacy of GIs, sample size, variance (R^2^), and effect size, we performed a power calculation using a non-centrality parameter framework^48^. For MR studies that did not describe R^2^, we used the extrapolated variance from other MR studies that used similar or identical genetic variants as instruments for calculation^20^. If there were meta-analyses for MR, we used the pooled effect estimates as the main outcomes rather than selecting a representative MR (Figure 3), and reviewed whether the meta-analyses of MR were properly conducted^49–53^.

**Fig. 3.**
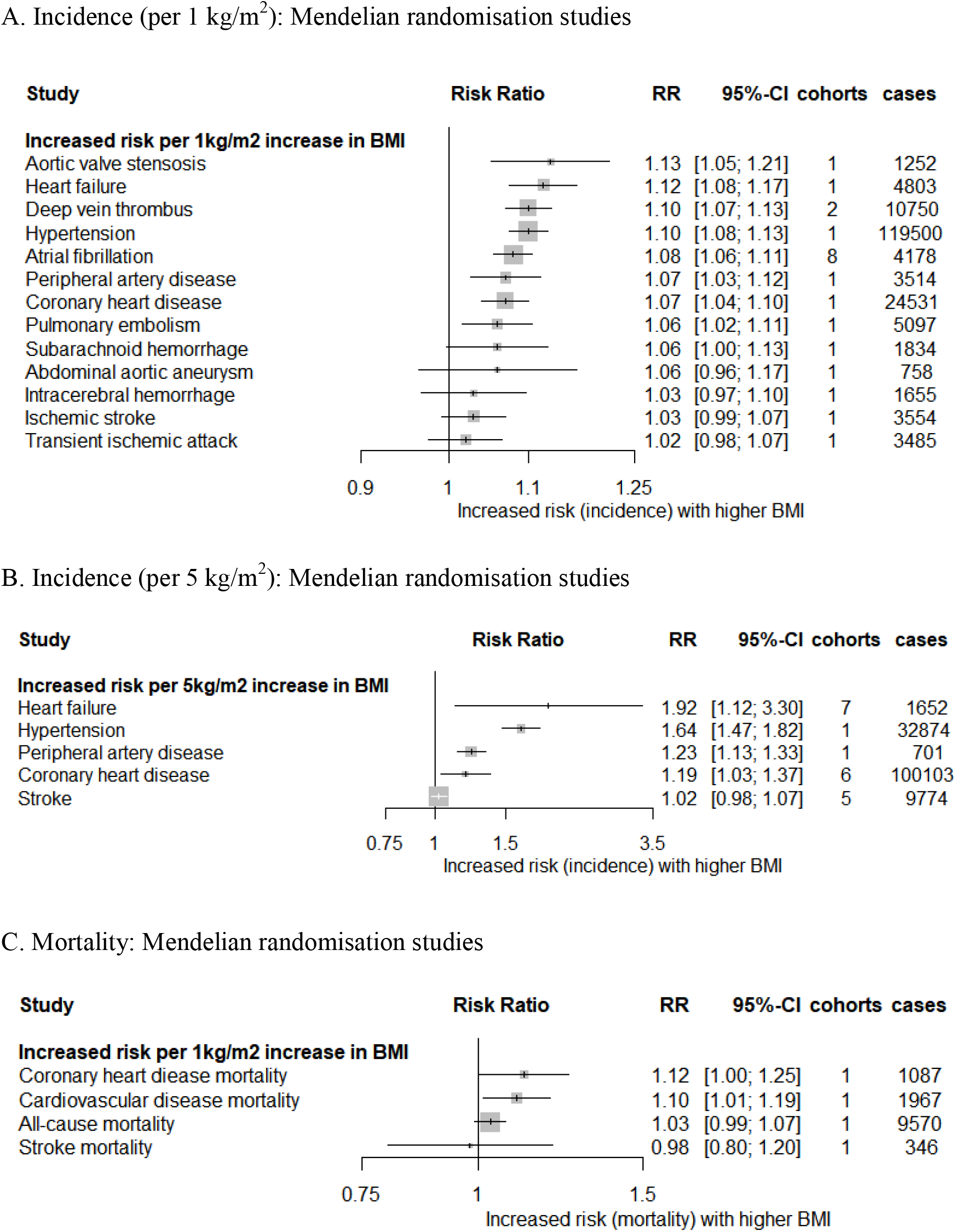
Collective results from Mendelian randomisation studies. (A) Increased risk of cardiovascular events per 1 kg/m^2^ increase in BMI. (B) Increased risk of cardiovascular events per 5 kg/m^2^ increase in BMI. (C) Increased risk of death per 1 kg/m^2^ increase in BMI. All results are based on random-effects models. The cohort and cases columns display the number of independent cohorts and the number of cases incorporated in the meta-analysis for the outcome. BMI, Body mass index; RR, Risk ratio or relative risk

### Evaluation of the certainty of evidence

We assessed the certainty of evidence for all reported associations using the Grading of Recommendations Assessment, Development, and Evaluation (GRADE) framework^54^, as has been done in numerous previous umbrella reviews^34 36 37 55^. The GRADE framework accounts for study limitations, imprecision, indirectness, inconsistency, publication bias, large magnitude of effect, and dose-response associations (Supplementary Table 4). The GRADE working group recommends assessing studies as grade 4 (high) for RCTs and grade 2 (low) for cohort studies. However, as RCTs are rarely possible for epidemiological topics, such as obesity, and large prospective cohort studies usually have the highest level of evidence^56 57^, we assigned grade 3 (moderate) for large-scale prospective cohort studies and grade 2 for retrospective cohort studies to account for such characteristics of epidemiological research. This modification to the GRADE approach is justified by numerous previous studies that have suggested a differentiation of evidence levels between prospective and retrospective cohort studies^56 58 59^. Imprecision was judged when the sample size was insufficient (< 1000) or when the CI was substantially large. When the CI value crossed 1, a judgement on imprecision was made according to our previous report^60^; if the CI value did not cross 1, we judged imprecision when the full CI length was wider than 0.5. Indirectness reflects differences in study populations; inconsistency was assigned when heterogeneity measured by the I^2^ statistic was greater than 50% for binary outcomes, and greater than 75% for continuous outcomes, since continuous outcomes are prone to higher heterogeneity than binary outcomes^61^, and the Cochrane Collaboration has a revised cut-off of I^2^ > 75% for high heterogeneity^32^. Publication bias was detected when the funnel plot was asymmetrical and the P-value for Egger’s test was < 0.10. A large magnitude of effect was considered when RR values were > 2 or < 0.5. A dose-response association was determined for the effect size that showed a proportional increase with adiposity indices. The final GRADE was determined after summation of all accountable biases. The 53 previously reported associations were classified according to the GRADE framework and are presented as evidence maps (Tables 1 and 2).

**Table 1.**
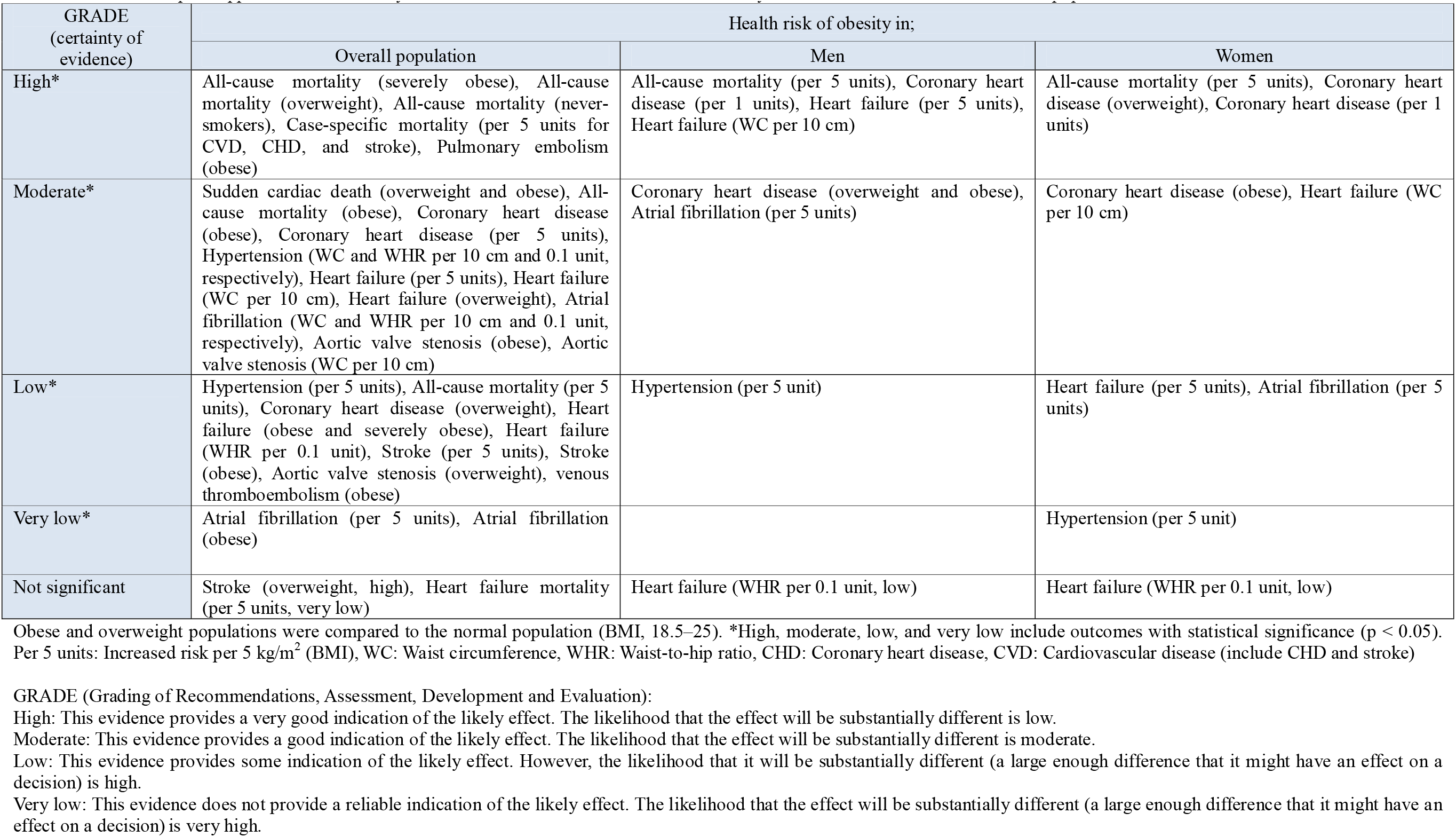
Evidence map for appraisal of the certainty of evidence for the association between obesity and cardiovascular outcomes in each population.

**Table 2.** Evidence map for appraisal of the certainty of evidence for the association between obesity and each cardiovascular outcome.

| Main outcomes | BMI categorical | BMI continuous<br>(per 5 kg/m<sup>2</sup>) | WC | WHR | Concordance<br>(between observational<br>study and MR) | Summary of evidence |
| --- | --- | --- | --- | --- | --- | --- |
| **Mortality** |  |  |  |  |  |  |
| All-cause mortality | High for overweight<br>Moderate for obese<br>High for severely obese | High for non-smokers<br>Low for all population | - | - | No (not significant in MR) | High in general, without causality supported by MR. Requires cautious interpretation |
| Heart failure | - | Not significant (very low) | - | - | MR not reported | Very low |
| Coronary heart disease | - | High | - | - | Yes | High, with causality supported by MR |
| Cardiovascular disease | - | High | - | - | Yes | High, with causality supported by MR |
| Stroke | - | High | - | - | No (not significant in MR) | High, without causality supported by MR. Requires cautious interpretation |
| Sudden cardiac death | Moderate for overweight<br>Moderate for obese | - | - | - | MR not reported | Moderate |
| **Incidence** |  |  |  |  |  |  |
| Coronary heart disease | Low for overweight<br>Moderate for obese | Moderate | - | - | Yes | Moderate in general, with causality supported by MR |
| Stroke | Not significant for overweight (high)<br>Low for obese | Low | - | - | Partially yes‡<br>(not significant in MR) | Low, without causality supported by MR |
| Heart failure | Moderate for overweight<br>Low for obese<br>Low for severely obese | Moderate | Moderate | Low | Yes | Low to moderate, with causality supported by MR |
| Atrial fibrillation | Very low for obese | Very low | Moderate | Moderate | Yes | Very low for BMI and moderate for WC & WHR, with causality supported by MR |
| Aortic valve stenosis | Low for overweight<br>Moderate for obese | - | Moderate | - | Yes | Moderate in general, with causality supported by MR |
| Hypertension | - | Low | Moderate | Moderate | Yes | Moderate in general, with causality supported by MR |
| Pulmonary embolism | High for obese | - | - | - | Yes | High, with causality supported by MR |
| Venous thromboembolism | Low for obese | - | - | - | Yes | Low, with causality supported by MR |

\*High, moderate, low, and very low include outcomes with statistical significance; ‡The association between BMI and the incidence of stroke in overweight populations reported in observational studies and the incidence of stroke reported in Mendelian randomisation studies are consistently not significant. BMI: Body mass index, WC: Waist circumference, WHR: Waist-to-hip ratio, MR: Mendelian randomisation study, CHD: Coronary heart disease, CVD: Cardiovascular disease (including CHD and stroke)

Although the GRADE framework is effective for assessing risk of biases, it does not account for reverse causation bias, which can be a major confounding factor in our topic of research^62^. Hence, we incorporated MR studies, and reviewed any discordance between observational studies and MR analyses for proper interpretation of the results, as summarised in Table 2.

Evidence from individual MR studies was assessed with statistical power (> 80%)^63^. The quality of each systematic review and meta-analysis was assessed using AMSTAR2^64^, as shown in Supplementary Table 5. We did not examine the quality of the individual cohort studies, since this was conducted by the authors of the included meta-analyses and was beyond the scope of this umbrella review^20^.

### Patient involvement

Neither the patients nor the general public were involved in the design, conduct, and reporting of the present study. No patient was asked to advise on interpretation or writing of the results. The results will be disseminated to the general public through public presentations and the authors’ involvement in different charities.

## RESULTS

### Literature review

Of the 16112 studies identified in the reviewed databases, 1322 were eligible for title and abstract review. After excluding 1001 studies that met our pre-specified exclusion criteria, 274 systematic reviews with meta-analyses and 47 MR studies were selected for full-text review. After full-text review, 283 studies were further excluded, and 11 systematic reviews with 53 meta-analyses, including a total of 448 cohort studies and 27 MR studies, were included for final analyses. The process of search and selection is presented in Figure 1. The AMSTAR2 grade was moderate for ten systematic reviews, while that for one review was graded as low (Supplementary Table 5).

### Meta-analyses of observational studies

The 53 meta-analyses of observational studies pooled from 448 cohorts were examined, 35 of which were added during our re-analysis. The reported analyses were categorised into 14 primary outcomes according to the disease entity: six morality outcomes, including all-cause mortality, CVD mortality, coronary heart disease (CHD) mortality, stroke mortality, heart failure death, and sudden cardiac death; and eight incidence outcomes, including CHD, stroke, heart failure, atrial fibrillation, aortic valve stenosis, hypertension, pulmonary embolism, and venous thromboembolism (Table 2).

All but four out of the 53 associations were statistically significant based on the results of the random-effect model (Table 1). The increase in the risk of developing CVD for every 5 kg/m^2^ increase in BMI ranged from 7% (RR, 1.07; 95% CI, 1.03 to 1.11) for stroke to 49% (RR, 1.49; 95% CI, 1.40 to 1.58) for hypertension (Figure 2A). The increase in the risk of developing CVD in the overweight population (BMI > 25 to 30 kg/m^2^) than in the reference group with normal BMI ranged from 14% (RR, 1.14; 95% CI, 1.10 to 1.18) for CHD to 38% (RR, 1.38; 95% CI, 1.14 to 1.68) for aortic valve stenosis. The increase in the risk of developing CVD in the obese population (BMI > 30 kg/m^2^) than in the reference group ranged from 16% (RR, 1.16; 95% CI, 1.06 to 1.26) for stroke to 124% (RR, 2.24; 95% CI, 1.92 to 2.61) for pulmonary embolism. The increase in the risk of mortality due to CVD for every 5 kg/m^2^ increase in BMI ranged from 5% (RR, 1.05; 95% CI, 1.04 to 1.07) for all-cause mortality to 49% (RR, 1.49; 95% CI, 1.45 to 1.53) for CVD mortality (Figure 2B).

The association of CVD outcomes with other adiposity measures, including WC and WHR, demonstrated consistent results with that of BMI (Supplementary Figure 1). The increase in the risk of developing CVD for every 10 cm increase in WC ranged from 18% (RR, 1.18; 95% CI, 1.12 to 1.25) for atrial fibrillation to 54% (RR, 1.54; 95% CI, 1.31 to 1.81) for aortic valve stenosis (Supplementary Figure 1). The increase in the risk of developing CVD for every 0.1 unit increase in WHR ranged from 8% (RR, 1.08; 95% CI, 1.01 to 1.15) for atrial fibrillation to 29% (RR, 1.29; 95% CI, 1.13 to 1.47) for heart failure (Supplementary Figure 1).

### MR studies

A total of 27 MR analyses were identified and classified into 22 outcomes, as presented in Supplementary Table 3. The proportion of variance (R^2^) explained by GI was 1.6% to 1.82%. Thirteen of the 22 outcomes were supported by a statistical power of greater than 80%. The increase in the risk of developing CVD for every 1 kg/m^2^ increase in BMI from MR analyses ranged from 6% (RR, 1.06; 95% CI, 1.02 to 1.11) for pulmonary embolism to 13% (RR, 1.13; 95% CI, 1.05 to 1.21) for aortic valve stenosis (Figure 3A). The increase in the risk of developing CVD for every 5 kg/m^2^ increase in BMI from MR analyses ranged from 19% (RR, 1.19; 95% CI, 1.03 to 1.37) for CHD to 92% (RR, 1.92; 95% CI, 1.12 to 3.30) for heart failure (Figure 3B). The increase in the risk of dying from CVD for every 1 kg/m^2^ increase in BMI from MR analyses ranged from 10% (RR, 1.10; 95% CI, 1.01 to 1.19) for CVD to 12% (RR, 1.12; 95% CI, 1.00 to 1.25) for CHD (Figure 3C).

### Subgroup analyses

The risk of CVD outcomes showed a proportional and dose-dependent increase with a step-up in BMI categories (Figure 4A). Obese men were more prone to unfavourable CVD outcomes than obese women (Figure 4B).

**Fig. 4.**
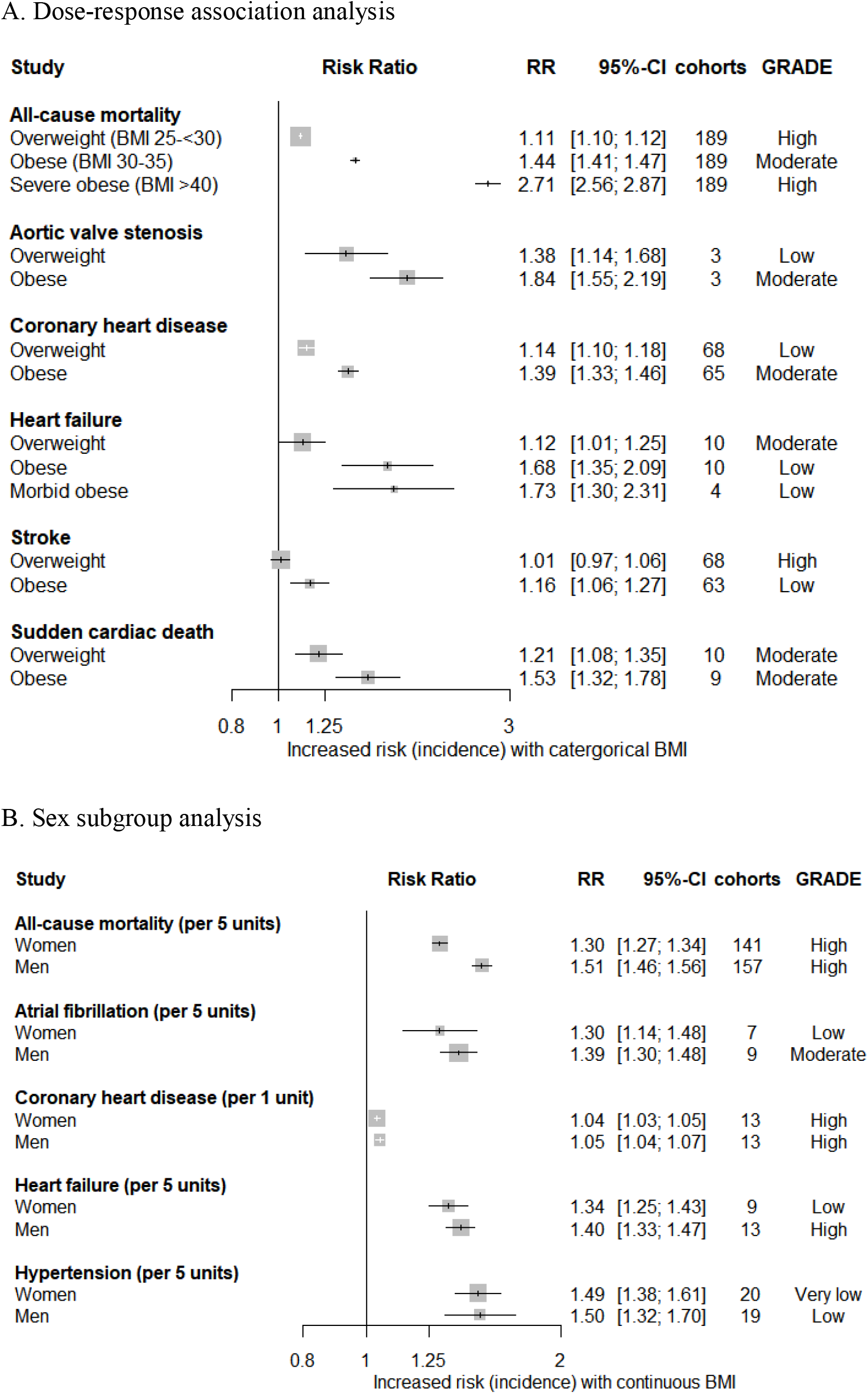
(A) Dose-response relationship of the incidence of cardiovascular events with step-up in categorical BMI. (B) Subgroup analysis for sex. All results are based on random-effects models. The cohort and participant columns display the number of independent cohorts and the total number of participants incorporated in the meta-analysis for the outcome. The certainty of evidence was evaluated with the GRADE framework. GRADE, Grading of Recommendations Assessment, Development, and Evaluation; BMI, Body mass index; RR, Risk ratio or relative risk

### Level of evidence

Of the 53 meta-analyses that investigated the effect of obesity on CVD-related outcomes, 15 associations (28%) were supported by high evidence certainty (GRADE), as described in the evidence map (Table 1); these meta-analyses summarised the data for overall population (n = 8), men (n = 4), and women (n = 3). A total of 19, 15, and 4 associations were supported by moderate, low, and very low certainty of evidence, respectively. All of the biases that we used in appraising the certainty of the grade are described in Supplementary Table 4: inconsistency (52%, 28/53), indirectness (0%, 0/53), imprecision (19%, 10/53), publication bias (21%, 11/53), large magnitude of effect (4%, 2/53), and dose-response association (79%, 42/53). To avoid reverse causation bias, concordance between the results of the observational and MR analyses, in either the direction and/or the statistical significance of associations, was identified and has been summarised in Table 2. The results of this study corroborate the causative effect of obesity on 9 of the 14 CVD-related outcomes; 4 mortality outcomes (all-cause mortality, heart failure death, stroke mortality, and sudden cardiac death), and the incidence of stroke had a risk of potential reverse causation bias.

For all outcomes, the shape of the p-curve (the distribution of statistically significant P-values for a set of findings, with right-skewed p-curves suggesting findings that contain evidentiary value) was significantly right-skewed (P < 0.05), indicating no evidence of p-hacking.

## DISCUSSION

This umbrella review paints a comprehensive picture of the existing evidence on the association between obesity and CVD by stratifying each association of obesity and CVD outcomes into distinct evidence levels. Eleven systematic reviews, with 53 meta-analyses, comprising a total of 488 cohorts and over 30 million participants were included herein for quantitative synthesis and quality assessment. As observational studies can suggest an association, but are unable to make claims on causation, MR studies were included to determine causality. Therefore, we provide results from observational studies and MR studies in parallel to contextualise both the magnitude of association and the direction of causality.

### Principal findings

Of the 53 meta-analyses that investigated the effect of obesity on CVD-related outcomes, 15 of the reported associations (28%) are supported by high evidence certainty (Table 1). While other reported associations might be genuine, various degrees of uncertainty remain. The causative effect of obesity on 9 out of the 14 CVD-related outcomes was corroborated by the results of this investigation; however, 4 mortality outcomes (all-cause mortality, heart failure death, stroke mortality, and sudden cardiac death) and the incidence of stroke have a risk of potential reverse causation bias (Table 2).

An increase in BMI was associated with a higher risk of developing all specific CVDs; incidence of CHD, heart failure, atrial fibrillation, stroke, hypertension, aortic valve stenosis, pulmonary embolism, and venous thromboembolism were all shown to increase with higher BMI, and the results were consistent in MR studies, with the exception of stroke (Figures 2 and 3). In our subgroup analyses, the risk of developing CVD showed a proportional and dose-dependent increase with a step-up in BMI categories, and obese men were more prone to unfavourable CVD outcomes than obese women (Figure 4). Association of CVD outcomes with other adiposity measures, including WC and WHR, demonstrated results that are consistent with those for BMI (Supplementary Figure 1).

Increased risk of all-cause mortality and CVD-specific mortality due to adiposity was supported by a high level of evidence from observational studies, but only the association between obesity and CHD mortality was shown to be statistically significant in MR studies (Table 2).

### Possible explanations

For MR analysis, Wade et al. used the polygenic risk score (PRS), comprising 77 single nucleotide polymorphisms associated with BMI as reported in the Genetic Investigation of Anthropometric Traits consortium, as the genetic instrument^65^, and the explanatory power of PRS for obesity was found to be reliable^66^. While there are still limitations to the MR approach^67, 68^, it is likely that potential biases are less marked than those of observational studies^65^ as long as the general assumptions for MR are met^69^. Triangulation of different methodologies is essential for inferring definite conclusions with proper causal inference^65^, and the findings from MR studies may add to the current body of evidence implicating obesity as a risk factor for cardiovascular health outcomes.

Of note, all-cause mortality significantly increased with higher BMI in observational analyses, but this relationship was not significant in MR analyses. Such discordance may be explained by the intrinsic limitation of observational studies in managing living confounders. Although a significant association of obesity with all-cause mortality rate was observed in 200 collective cohorts, with adjustment for age, sex, and smoking, this association should be interpreted cautiously as all-cause mortality may involve diverse causes of death, such as pneumonia and trauma, which may have weaker links to obesity. Considering that CVD-related mortality was significantly high with increased obesity in both observational and MR analyses (Table 2), it is plausible that the observational results of all-cause mortality may have been overestimated by other intermediate or surrogate causes for death in cohorts that are not driven by cardiovascular impairment.

The risk of the incidence of all CVDs, except stroke, was significantly increased with obesity in both observational and MR analyses (Figures 2 and 3). A large number of mediators released by adipose tissue may play key roles in the link between obesity and CVD; adipose tissues release bioactive mediators that influence alterations in lipids, coagulation, fibrinolysis, and inflammation, leading to endothelial dysfunction and atherosclerosis^70^. Atherosclerosis is the principal origin of CVD^71, 72^, and synergistically interacts with hypertension, and both factors aggravate one another^71, 73^. It is notable that hypertension was the most vulnerable entity affected by BMI in our analysis; the increase in the risk of developing CVD for every 5 kg/m^2^ increase in BMI ranged from 7% (RR, 1.07; 95% CI, 1.03 to 1.11) for stroke to 49% (RR, 1.49; 95% CI, 1.40 to 1.58) for hypertension. Other CVDs may be the consequences of atherosclerosis and hypertension, as these entities represent pathophysiological basis and are thus major risk factors for CVD^72 74–77^.

Several MR studies have reported that obesity has no causal effect on stroke^78–80^, and stroke was the least affected entity in our observational analysis (Figure 2). This result is in line with a recent study conducted by Khera et al. in which stroke occurred less frequently than most of other CVDs, such as hypertension and venous thromboembolism, in high BMI-PRS carriers (10^th^ percentile)^66^; this observation probably indicates that the genetic drivers for obesity have a weak causal effect on the development of stroke. The discordance in the results of observational and MR analyses for stroke in our umbrella review may suggest that stroke pathophysiology involves a complex mechanism in which obesity is only a minor part^81^.

### Clinical implications and future research

Obesity is a multifactorial disease that results from interactions between genetics and lifestyle^82 83^. The heritability for obesity is known to be around 40%^82 84^, and the remainder could be explained by lifestyle factors, which suggests obesity as a modifiable risk factor^85^. In this context, the causal effect of obesity on nearly all specific cardiovascular outcomes suggested in this study provides an enthusiastic prospect in which lifestyle modification to reduce adiposity can result in the overall reduction of cardiovascular health problems and substantial health-economic burden^86^. This study supports the assertions that reducing adiposity through interventional approaches, such as bariatric surgery^87^; promotion of educational equality^88^; lifestyle modifications, including healthy diets^89–94^; and increasing physical activities^93^ may provide a better chance of improving one’s well-being^95^, even more so than previously expected, by affecting multiple vascular health outcomes. Our results also support the diet and lifestyle recommendation proposed by the American Heart Association (AHA)^96^, and further specify the benefits. Future studies should be conducted to provide empirical evidence of the effect of lifestyle modifications targeted at reducing adiposity on cardiovascular benefits.

### Limitations

This study has several limitations. First, umbrella reviews have intrinsic limitations in that they only focus on existing meta-analyses, and therefore important phenotypes that were not assessed in a meta-analysis level may be overlooked. To minimise the disregard of clinically relevant cardiovascular phenotypes, we independently conducted de-novo meta-analyses for certain CVDs (e.g., aortic valve stenosis) that have not been meta-analysed despite a sufficient number of published original studies. Second, when meta-analyses are outdated, they may provide incomplete conclusions with less power, which may directly affect the analyses of subsequent umbrella reviews. As a countermeasure, we updated 19 meta-analyses of observational studies by incorporating recent reports from 35 cohorts to reflect up-to-date conclusions. Third, we did not analyse the effect of being underweight on CVD outcomes. Although lower BMI is known to affect CVD outcomes and constitutes a J-shape curve, this analysis was beyond the scope of this investigation, since our research question mainly lies in how increased adiposity affects CVD outcomes. Although our study is similar to previous umbrella reviews on obesity^1, 3^, none of these studies included being underweight in their analyses. Lastly, the value of RR should be interpreted with caution as the equivalent RR was retrieved based on assumptions and approximations. We approximated different metrics to equivalent RRs using Fusar-Poli et al.’s guideline^43^, and adapted an approach of coordinating the results with different metrics used in previous meta-analyses^44, 45^ and umbrella reviews^20,46^; nevertheless, the equivalent RR may not reflect the actual effect size, and rather should be taken as inferences that allow comparisons of the magnitude of the effect of obesity on different cardiovascular phenotypes and outcomes.

### Conclusion

Although obesity as a risk factor for various cardiovascular outcomes has been extensively studied for decades, only 15 of the 53 reported associations (28%) were supported with a high evidence level. While other associations could be genuine, various degrees of uncertainty remain. The causative effect of obesity on 9 out of 14 CVD-related outcomes was corroborated by the results of this study; 4 mortality outcomes and the incidence of stroke remained at risk of potential reverse causation bias and requires further clarification. Since obesity is progressively increasing around the globe, and CVD is continually threatening public health, understanding the gradient of evidence behind the associations between these two clinical entities is necessary. The strong associations discovered between obesity and CVD outcomes in this review should be considered in clinical practice and in the formulation of health policies, while any weak links should be reinforced with further scientific research.

### Funding

This work was supported by the National Research Foundation of Korea (NRF) grant funded by the Korea government (MSIT) (No. 2019R1A2C4070496). MS Kim had full access to all the data in the study, and HH Won made the final decision to submit for publication.

### Contributors

MS Kim contributed to the concept and design of the study. MS Kim and WJ Kim identified and acquired relevant trials and extracted data. MS Kim drafted the protocol for this study. MS Kim analysed and interpreted the data. MS Kim wrote the first draft of the manuscript. HH Won and AV Khera contributed to the interpretation of the data and the critical revision of the manuscript. All authors read and approved the final submitted version.

## Data Availability

Not applicable

## Ethical approval

Not required.

## Data sharing

Not applicable.

## Dissemination declaration

We plan to disseminate the results to study participants and or patient organisations

## Transparency

The manuscript’s guarantors (HH Won) affirm that this manuscript is an honest, accurate, and transparent account of study being reported, that no important aspects of the study have been omitted, and that any discrepancies from the study as planned have been explained.

